# SARS-CoV-2 positivity in asymptomatic-screened dental patients

**DOI:** 10.1101/2020.12.30.20248603

**Authors:** DI Conway, S Culshaw, M Edwards, C Clark, C Watling, C Robertson, R Braid, E O’Keefe, N McGoldrick, J Burns, S Provan, H VanSteenhouse, J Hay, R Gunson, Dental COVID-19 Surveillance Survey Group

**Affiliations:** School of Medicine, Dentistry, Nursing, University of Glasgow, Glasgow, UK; Public Health Scotland, Glasgow, UK; Department of Public Health, NHS Ayrshire and Arran, Ayr, UK; Mathematics and Statistics, Strathclyde University, Glasgow, UK; Public Health Department, NHS Fife, Leven, UK; Oral Health Directorate, NHS Greater Glasgow & Clyde, Glasgow, UK; Lighthouse Lab in Glasgow, Glasgow, UK; BioClavis, Glasgow, UK; West of Scotland Specialist Virology Centre, Glasgow Royal Infirmary, Glasgow, UK

**Keywords:** COVID-19, Severe Acute Respiratory Syndrome Coronavirus 2, Asymptomatic, Dentistry, Outpatient, Swab

## Abstract

Enhanced community surveillance is a key pillar of the public health response to COVID-19. Asymptomatic carriage of SARS-CoV-2 is a potentially significant source of transmission, yet remains relatively poorly understood. Disruption of dental services continues with significantly reduced capacity. Ongoing precautions include pre- and/or at appointment COVID-19 symptom screening and use of enhanced personal protective equipment (PPE). This study aimed to investigate SARS-CoV-2 infection in dental patients to inform community surveillance and improve understanding of risks in the dental setting. Thirty-one dental care centres across Scotland invited asymptomatic screened patients over 5-years-old to participate. Following verbal consent and completion of sociodemographic and symptom history questionnaire, trained dental teams took a combined oropharyngeal and nasal swab sample using standardised VTM-containing testkits. Samples were processed by the Lighthouse Lab and patients informed of their results by SMS/e-mail with appropriate self-isolation guidance in the event of a positive test. Over a 13-week period (from 3August to 31October2020) n=4,032 patients, largely representative of the population, were tested. Of these n=22 (0.5%; 95%CI 0.5%, 0.8%) tested positive for SARS-CoV-2. The positivity rate increased over the period, commensurate with uptick in community prevalence identified across all national testing monitoring data streams. All positive cases were successfully followed up by the national contact tracing program. To the best of our knowledge this is the first report of a COVID-19 testing survey in asymptomatic-screened patients presenting in a dental setting. The positivity rate in this patient group reflects the underlying prevalence in community at the time. These data are a salient reminder, particularly when community infection levels are rising, of the importance of appropriate ongoing Infection Prevention Control and PPE vigilance, which is relevant as healthcare team fatigue increases as the pandemic continues. Dental settings are a valuable location for public health surveillance.

## Introduction

In less than a year, the coronavirus disease (COVID-19) pandemic has wreaked a devastating global impact – causing harm to people’s health, society, and the economy[**1**].

Trusted and reliable surveillance and epidemiology programs providing robust data are central to the efforts to monitor, understand, and respond to the pandemic – globally these include the World Health Organisation[**1**], the Johns Hopkins Coronavirus Resource Centre[**2**] and in Scotland there are Public Health Scotland (PHS) resources[**3**].

From 2019 December 31, the World Health Organisation (WHO) was alerted of an outbreak of “pneumonia of unknown cause” in Wuhan, China[**1**]. The virus causing the outbreak was later formerly named severe acute respiratory syndrome coronavirus 2 (SARS-CoV-2). The WHO declared COVID-19 a pandemic on 11 March[**1**]. At the time of writing (2020 November 16) globally, there were over 54 million cases and over 1.3 million deaths estimated[**2**].

Asymptomatic carriage of SARS-CoV-2 is a potentially significant source of transmission, yet remains relatively poorly understood[**4**]. There are wide-ranging estimates of the level of asymptomatic SARS-CoV-2 infection, which vary by setting, country, and over time[**5**]. The reported proportions of individuals testing positive who are asymptomatic have ranged from: as low as 5% in a small hospital study from China in the early stages of the pandemic[**6**]; to 17.9% in the cruise ship in Japan in February[**7**]; to 42.5% from the village-wide study in Italy during the first wave lockdown[**8**]; and to 76.5% of those tested positive in these preliminary Office for National Statistics (ONS) population-level data from England[**9**]. Recent systematic reviews and meta-analyses estimate the prevalence of asymptomatic SARS-CoV-2 infection to be less than 20%. In July 2020, a review of 41 studies (n=50,155 participants) found a pooled percentage of 15.6% (95%Confidence Interval CI 10.1%, 23.0%) but with significant heterogeneity and a very wide range[**10**]. In September 2020, 94 studies (n=25,538 participants) estimated 20% (95%CI 17%, 25%) asymptomatic carriage, suggesting that inflated results from studies earlier on the pandemic were due to a limited range of symptoms being included[**11**]. More recently, in October 2020, a focused meta-analysis of 13 low risk of bias studies (n=21,708 participants) found 17% (95%CI 14%, 20%) with a range of 4 to 41%[**12**]. These reviews identified biases due to selection of study participants, calling for future population representative studies to determine the true proportion of asymptomatic SARS-CoV-2 infections as well as the prevalence in the population. Incomplete symptom assessment has also been considered important in overestimating the asymptomatic fraction[**13**].

Due to the risks of transmission associated with receiving dental care, during the lockdown period in Scotland, all National Health Service dental practices were not able to see patients on their premises. Across Scotland over seventy Urgent Dental Care Centres (UDCCs) were established for the provision of emergency dental treatment. An opportunity for dentistry to contribute to the COVID-19 testing and (asymptomatic) surveillance effort in Scotland was proposed on the basis that patients were already making an essential visit outside lockdown; that dental teams could accurately screen patients to ensure they had no COVID-19 symptoms, and could readily be trained to undertake COVID-19 swab testing. Dental teams would also already be wearing appropriate personal protective equipment (PPE) – which would both maximise PPE use and save the need for additional PPE given shortages at the time. Moreover, NHS dental settings in Scotland had previously been shown to be a feasible source of participants representative of the population for virus (HPV) testing[**14**].

This study aimed to investigate SARS-CoV-2 infection in asymptomatic-screened dental patients to inform community surveillance and improve understanding of risks in the dental setting.

## Materials and Methods

### Protocol

A protocol for testing asymptomatic patients attending dental settings was developed by adopting and adapting the primary care community surveillance program in Scotland where patients with suspected COVID-19 symptoms were being tested[**15**]. A National Steering Group for the Dental COVID-19 Surveillance Program was established. The program was piloted in two Health Boards (Fife and Greater Glasgow & Clyde) before being rolled out across Scotland. Initially patients were recruited only from Urgent Dental Care Centres. As dentistry began to remobilise, the program was expanded to clinics in hospital dental services, public dental services, and a small number of general dental practices.

### Training

A training video developed by Health Protection Scotland for healthcare professionals undertaking swab testing was made available to dental teams[**16**]. The National Steering

Group set up meetings with each Board prior to them commencing swabbing, to allow a “walk through” of procedures.

### Approvals

The West of Scotland NHS Research Ethics Service was approached and waived the requirement for research ethics approval – advising that the study: “falls within the definition of Usual Practice in Public Health, … Designed to investigate an outbreak or incident to help in disease control and prevention.” Information Governance approval was obtained via Public Health Scotland following completion of a Data Privacy Impact Assessment (DPIA; DP20210155).

### Inclusion criteria

All dental patients over 5-years of age attending selected dental settings across Scotland who were screened to be asymptomatic of COVID-19 symptoms on attendance and were eligible were invited to have a voluntary COVID-19 swab test, subject to clinician’s discretion. A patient information letter (available in several languages) was provided to patients prior to or on their arrival at the clinic[**17**]. Following surveillance protocols[**15**], the patients were further informed of the process and verbal consent was obtained, recorded in the clinical notes, and on the testkit registration portal[**18**].

### Data collection

A member of the dental used a questionnaire to obtain information from the patient. This documented sociodemographic information: forename, surname, date of birth, gender, postcode, Community Health Index (CHI number – the unique National Health Services NHS identifier in Scotland) and ethnicity; past medical history, previous/current history of potential COVID-19 symptoms (via a checklist: fever, cough, shortness of breath, fatigue, wheeze, headache, sore throat, altered smell/taste, vomiting, diarrhoea, upper / lower respiratory tract infection), self-reported “shielding” (the policy in Scotland where people were asked by their GP to stay at home and minimise all contact with others for 12 weeks [**19**]); and public health behaviours (mask wearing, physical distancing, home disinfection, extra handwashing and use of hand sanitiser).

The information from the questionnaire was used to populate the UK Government Department of Health and Social Care (DHSC) web-based COVID-test swab registration form[**18**], which collected: Unique Organisation Number of dental setting; patient details (forename, surname, date of birth, gender, ethnic group, country of residence, postcode, first line of address, CHI); note of patient having any coronavirus symptoms; testkit barcode, date and time of swab, patient email address and mobile phone number. There were two tick boxes to confirm that consent had been given to take the swab and enter the data.

### Swab testing

In the dental surgery, a combined oropharyngeal and nasal swab, using standardised VTM-containing (“non-Randox”) testkits, was taken by an appropriately trained member of the dental team. The swabs were transported to the Lighthouse Lab in Glasgow (LliG), one of the network set up by the UK Government as part of their COVID-19 testing strategy[**20**].

### Laboratory methods

The testing system used at the Glasgow Lighthouse Lab consists of the ThermoFisher Scientific KingFisher Flex System Nucleic acid extraction, the TaqPath™ COVID19 CEIVD RTPCR Kit, a multiplex assay that targets three SARS-CoV-2 genomic regions (ORF1ab, S protein and N protein) and bacteriophage MS2 (internal control), and run on the Applied Biosystems™ 7500 Fast RealTime PCR Instrument (used with 7500 Software v2.3). Data were analysed using UgenTec Fast Finder 3.300.5 (TaqMan 2019-nCoV Assay Kit V2 UK NHS ABI 7500 v2.1). The Assay Plugin contains an Assay specific algorithm and decision mechanism that allows conversion of the qualitative amplification Assay PCR raw data from the ABI 7500 Fast into test results with minimal manual intervention. Samples are called positive in the presence of at least single N gene and/or ORF1ab but may be accompanied with S gene (1, 2 or 3 gene positives). S gene was not considered a reliable single gene positive.

The analytical sensitivity was determined to be 200 copies/ml by assessment of a Qnostics SARS-CoV-2 AQP linearity panel. The analytical specificity was assessed by ongoing participation in an external EQA scheme provided by QCMD (CVOP20S). The clinical sensitivity and specificity of the LLiG SARS-CoV-2 assay was determined by assessment of two clinical sample panels (positives and negatives). Whole system reproducibility/precision was determined by comparing Ct values over four days’ consecutive days (including different extractions, PCR platforms and operators).

### Data management

Patients were informed directly of their results (positive, negative, or void) by SMS/email. Positive test results entered the Electronic Communication of Surveillance in Scotland (ECOSS) system and the Case Management System (CMS) used by health protection teams in local Boards for contact tracing (known as the NHS Scotland’s “Test and Protect” system[**21**]). Patients with a positive result were followed up by local Test and Protect teams, and the National Steering Group contacted the relevant health protection team, consultant in dental public health and director of dentistry to ensure all were aware the test had been part of an asymptomatic surveillance program. Those patients with void (test “could not be read”) results were followed up by the National Steering Group, as the standard SMS message (intended primarily for symptomatic testing) recommended a retest, which was not required for an unclear asymptomatic surveillance result.

### Statistical analysis

Positivity percentages (rates) with 95% Confidence Intervals (CIs) were calculated using Wilson’s method (and 3-week rolling averages) were calculated on a weekly basis and incorporated into the PHS enhanced surveillance of COVID-19 in Scotland weekly report for Scottish Government. Rates and 95%CIs were similarly calculated for the 13-weeks overall (and also for the first 6-weeks and second 7-weeks). For data visualisation comparison, routine daily testing data for the same time period were extracted from the PHS daily positive test monitoring dashboard[**3**]. Growth rates were estimated using poisson regression models with a log link and an interaction test was used to compare the growth rate between our dental and General (Medical) Practice (GP) Primary Care Surveillance data[**15**].

Participant characteristic proportions and confidence intervals were calculated using Wilson’s method. These were compared against population demographics by linking our data into the Early Pandemic Evaluation and Enhanced Surveillance of COVID-19 (EAVE-II) linked dataset in PHS which has demographic and GP clinical information from the whole population of Scotland[**22**]. Population weightings were calculated via comparing the participants to the EAVE-II dataset on age group, socioeconomic deprivation, and co-morbid risk groups. Gender was not included in the calculation of the weights as the proportions of men and women in our dental surveillance sample matched the population proportions.

Analysis was undertaken to investigate the impact of using an imprecise test by assessing the impact on positivity by various scenarios of specificity and sensitivity. Very high levels of specificity were used in line with ONS (99.92%)[**23**] and PHS (99.68%) data (unpublished), alongside lower levels of sensitivity as in ONS[**23**], 60% (with 95% between 45% and 75%), or medium sensitivity of 90% (with 95% probability between 85% and 95%).This was carried out by a parametric bootstrap analysis with specificities and sensitivities coming from beta distributions and test positivity from a binomial. All analyses were undertaken in BusinessObjects, SPSS, and R version 3.6.1.

## Results

By week 13 of the program, 31 dental centres from 13 of the 14 Health Boards across Scotland were recruiting patients into the program. There were 4,032 tests processed during this period. Compared to the Scottish COVID-19 surveillance population distribution, the sample had similar sex distribution, slightly fewer participants from the least deprived communities, and fewer children (Table 1). The sample had similar numbers of recorded co-morbid conditions, but also had a high proportion (16.9%) of participants who self-reported that they had been shielding compared with the population numbers reported by PHS (n=179,728; 3.3% of the population)[**22**].

**Table 1.**
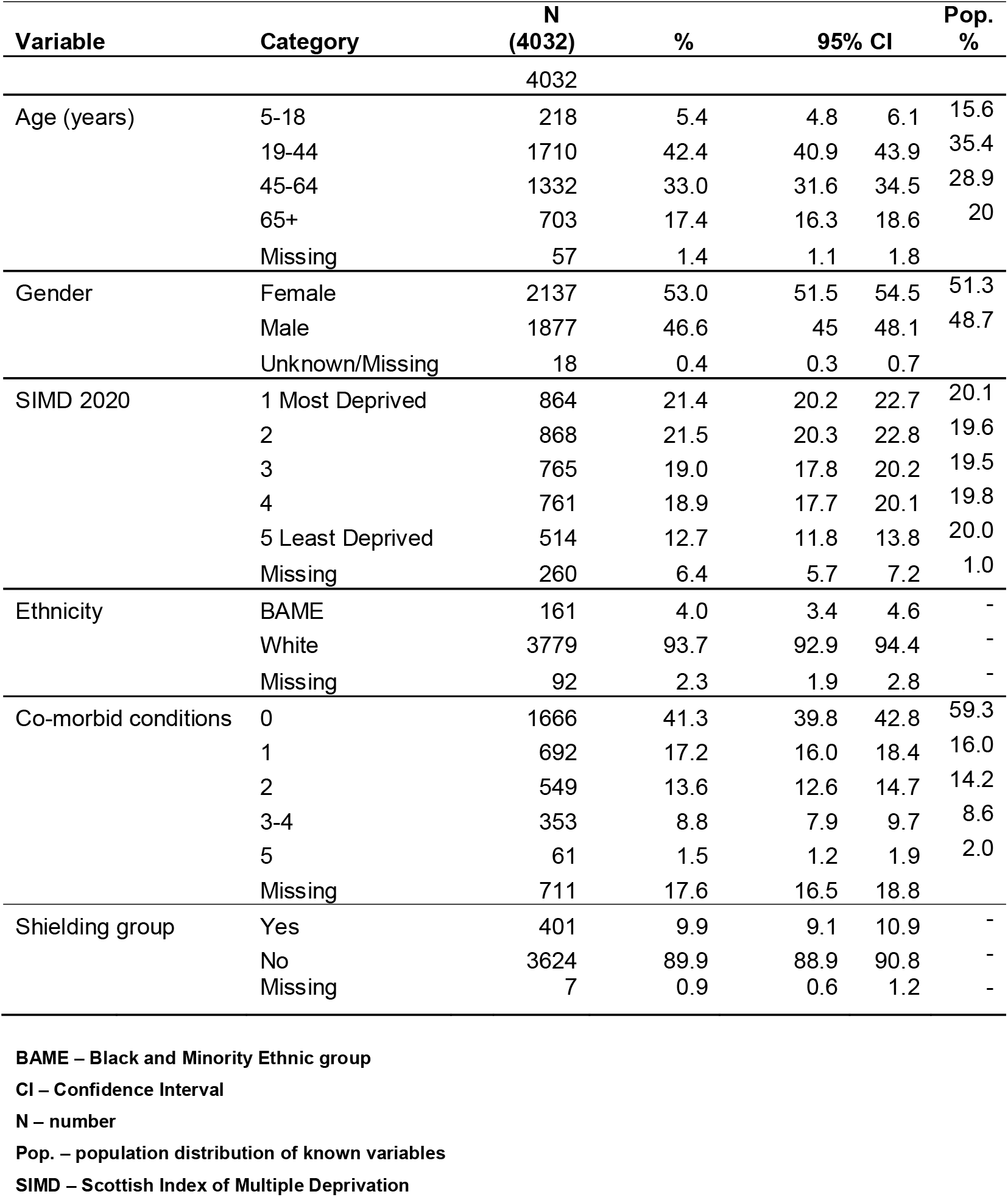
Characteristics of participant dental patients with swab test results.

There were 22 positive tests in total during the 13 weeks (Table 2, Cycle Thresholds CTs reported in Appendix Table 1), with an overall test positivity rate of 0.6% (95%CI 0.4%, 0.8%). None of the positive tests had the S-gene drop-out (Appendix Table 1) suggestive of the “new UK variant” referred to as SARS-CoV-2 VUI 202012/01. Analysis with population weighting had a minimal impact on the overall population percentage positive (0.6% 95%CI 0.4%, 0.9%). There was an increase from an average of 0.0% (95%CI 0.0%, 0.8%) during the first 6-weeks when no patients tested positive to 0.7% (95%CI 0.5%, 1.1%) in the latter 7-weeks (Table 2 and Figure 1). This trend followed the second wave uptick observed in the daily positive test data (Figure 2).

**Table 2.** **Numbers of samples, test results, and percentage positive for SARS-CoV-2 per week (by specimen date; dental settings; Scotland; 2020 August 2020 to 2020 October 31)**

| Week (Date) | No. samples | No. void tests | No. positive tests | % swab positive | 95% CI |  |
| --- | --- | --- | --- | --- | --- | --- |
| Week 1 (August3 -) | 72 | 1 | 0 | 0% | 0% | 5.1% |
| Week 2 | 109 | 1 | 0 | 0% | 0% | 3.4% |
| Week 3 | 87 | 3 | 0 | 0% | 0% | 4.2% |
| Week 4 | 85 | 0 | 0 | 0% | 0% | 4.3% |
| Week 5 | 317 | 43 | 0 | 0% | 0% | 1.2% |
| Week 6 | 334 | 55 | 0 | 0% | 0% | 1.1% |
| Week 7 | 390 | 16 | 2 | 0.5% | 0.1% | 1.9% |
| Week 8 | 397 | 7 | 0 | 0% | 0% | 1.0% |
| Week 9 | 398 | 2 | 1 | 0.3% | 0% | 1.4% |
| Week 10 | 419 | 13 | 7 | 1.7% | 0.8% | 3.4% |
| Week 11 | 432 | 5 | 1 | 0.2% | 0.0% | 1.3% |
| Week 12 | 510 | 9 | 6 | 1.2% | 0.5% | 2.5% |
| Week 13 (October26 -) | 482 | 15 | 5 | 1.0% | 0.4% | 2.4% |
| **Total (Weeks 1-6)** | **1004** | **103** | **0** | **0%** | **0%** | **0.4%** |
| **Total (Weeks 7-13)** | **3028** | **67** | **22** | **0.7%** | **0.5%** | **1.1%** |
| **Total (Weeks 1-13)** | **4032** | **170** | **22** | **0.6%** | **0.4%** | **0.8%** |

CI – Confidence Interval
No. – number

**Figure 1.**
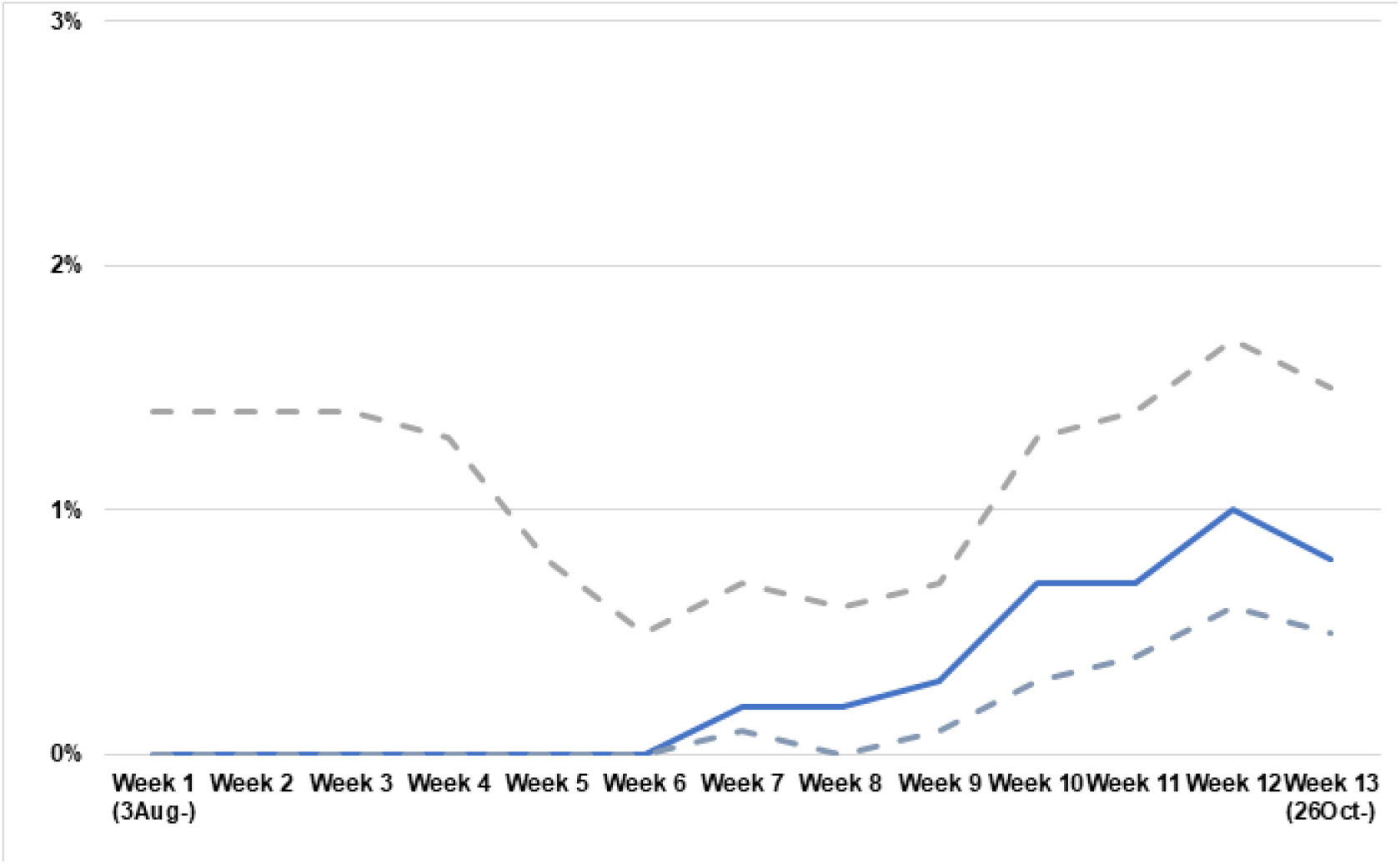
**Percentage positive tests (solid line) with upper and lower 95% Confidence Intervals (dashed lines) per 3-week average by specimen date; dental settings; Scotland; 2020 August 3 to 2020 October 31**

**Figure 2.**
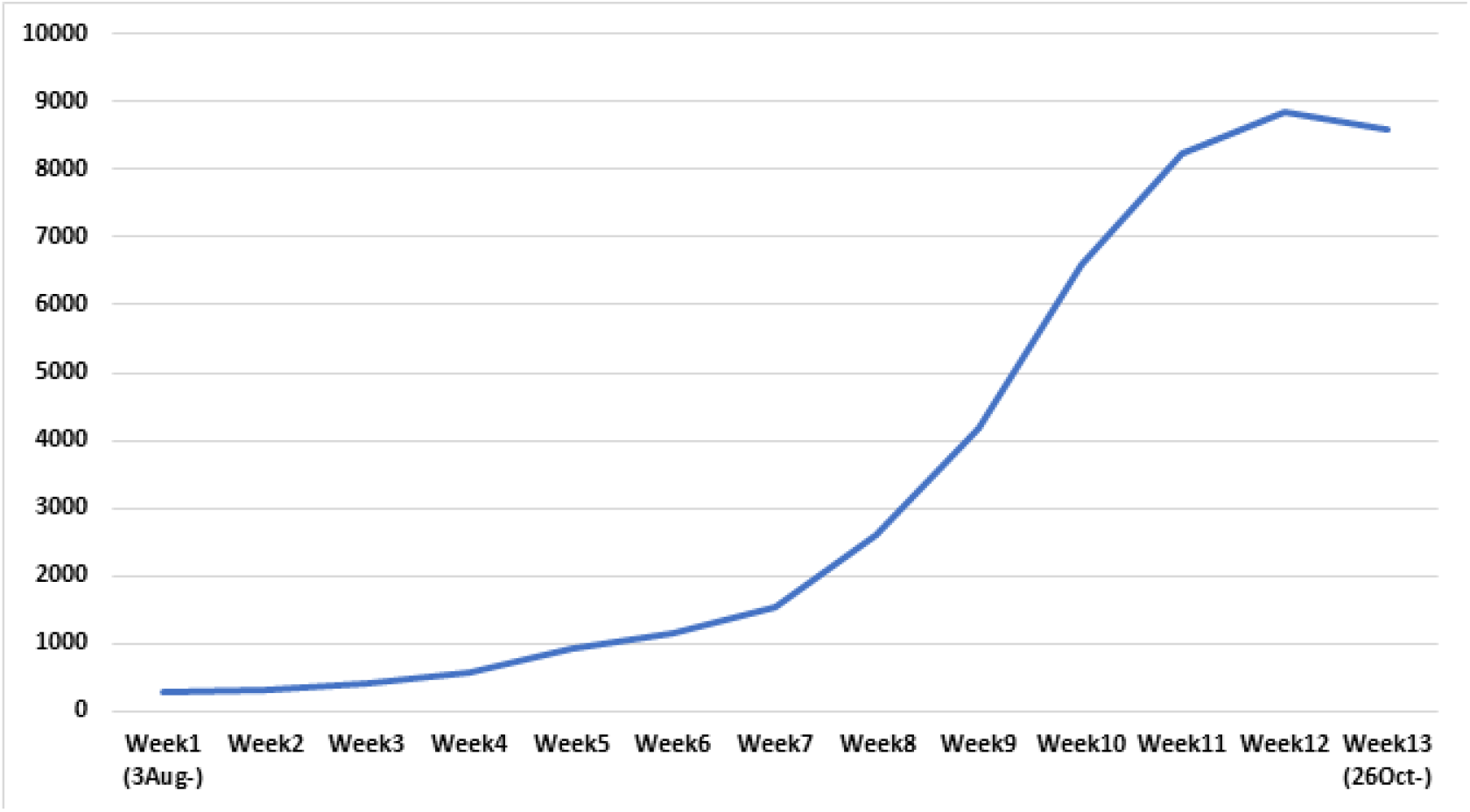
**Number of positive tests per week by specimen date; all settings Scotland; 3AUG2020 to 31OCT2020 – Public Health Scotland COVID-19 monitoring dashboard data[3]**

Over the period from week 30 to week 44 the estimated growth in the epidemic measured via the GP Primary Care Surveillance Program of symptomatic patients was 0.31 (95% CI 0.28%, 0.35%) per week (data not shown [**15, 22**]) and for the dental surveillance of asymptomatic individuals was 0.29 (95% CI 0.10%, 0.48%) per week. There was no difference in growth rates (p = 0.92), though the dental growth rate was estimated with lower precision due to the small numbers of positive cases.

We estimated the adjusted positivity for both a very high or high specificity each combined with moderate or low sensitivity. With a very high specificity (99.92%) and moderate sensitivity of 90% the adjusted overall positivity was 0.5% (0.2%, 0.8%). With a very high specificity (99.92%) and a low sensitivity of 60% the adjusted overall positivity increased to 0.8% (0.4% 1.4%). With a slightly lower specificity (99.68%) and moderate sensitivity (90%) the adjusted positivity rates were 0.3% (0.0%, 0.6%). And at the slightly lower specificity (99.68%) with low (60%) sensitivity the adjusted positivity was 0.4% (0.0%, 1.0%).

Of the tests which were classified “void” (n=170, 4.2%), the majority (n=108) occurred in a three-week (weeks 5-7) cluster (Table 2). At this time testing demand frequently exceeded laboratory processing capacity causing delays[**24**]. Indeed, analysis of the time between date of sample taken and date of processing in the lab was significantly (p<0.01) longer in the void samples (median 5.0 days, mean 4.2 days) compared with the processed (median 3.0 days, mean 3.1 days) test groups (data not shown).

## Discussion

To the best of our knowledge, this is the first report of SARS-CoV2 positivity in asymptomatic dental patients, and documentation that asymptomatic carriage in this population shows a pattern of epidemic growth consistent with the surveillance of symptomatic individuals[**3, 15, 22**].

Our results are similar to those from the Office for National Statistics (ONS) Coronavirus (COVID-19) Infection Survey which commenced in Scotland in October 2020 – a large household population-based study assessing the incidence of infection in the UK general population. Data are produced fortnightly and have shown a rise from 0.6% (95%CI 0.4%, 0.9%) at the beginning of October to 0.9% (95%CI 0.6%, 1.2%) by the end of the month[**25**]. The ONS survey has no symptom-related exclusion criteria and analyses from the pilot study in England found three-quarters (n=88/115) of the participants who tested positive for SARS-CoV-2 were asymptomatic among the total 36,061 tested (between April 26 and June 27)[**9**]. A more recently published systematic review[**26**] identified only two general population studies, which found the proportion of asymptomatic COVID-19 at the time of testing to be 20% in Luxembourg (n=1 out of the 5 positives from 1842 tested[**27**] and 75% in Italy (n=6 out of the 8 positives from 2322 tested[**8**]. In the same systematic review, they identified five small cohorts of obstetric patients from USA and Japan – the proportion of asymptomatic patients was between 84 and 100% in the smallest studies (none larger than n=155) and 45% in the biggest (20 out of 757 patients with positive tests).

There have been relatively few studies investigating SARS-CoV-2 infection in COVID-19 asymptomatic-screened patients in clinical outpatient healthcare (including dental) settings. One survey conducted in outpatient gastrointestinal clinics prior to endoscopy found three from 2611 (0.1%; 95%CI 0.0%, 0.3%) asymptomatic patients tested positive for SARS-CoV-2 on nasopharyngeal swab[**28**]. Most of the other clinical based testing studies have been undertaken in patients admitted to hospital, e.g. prior to surgery[**29**].

The estimates provided in our surveillance study are percentage of dental patients testing positive for the SARS-CoV-2 (the positivity rate). We were unable to report the population prevalence rate because without a true gold standard diagnostic test, we do not know the accurate sensitivity (true-positive rate) and specificity (true-negative rate). Given the low number of positive tests in the study, even if all the positives were false the specificity would be very high, confirmed in the ONS population pilot in England[**30**]. The test sensitivity has previously been estimated to be between 85% and 98%[**30**]. As underlying prevalence rises, high test specificity would not lead to changes in false positives; however, with low test sensitivity there would be an increase in the numbers of false negatives and the proportion of all positives that are false would also decrease[**31**]. We explored the potential impact of varying degrees of specificity and sensitivity. As our program uses the same Lighthouse Lab testing as the ONS survey, the most likely scenario takes their reported higher specificity level[**30**] alongside the moderate level of sensitivity (which is the most likely scenario as the underlying population prevalence has been rising over the survey period[**31**]). In this scenario we find our overall positivity rate unchanged.

The CT values show a significant number of positive tests, which were reported as “weak/close to the limit of detection”, while they are likely true analytical positives, the health protection teams would ordinarily risk assess and propose a repeat test. However, in practice, in the height of the pandemic, these positives (irrespective of symptoms) were taken on face value and all patients were advised to self-isolate and were followed up for contact tracing. Similarly, the sheer volume of testing from multiple locations and surveillance streams during the pandemic caused lab capacity and delay issues in processing, which led to some test results being void.

This surveillance program had several advantages including using trained dental healthcare teams for the collection of high quality and complete data and samples, although supervised self-collected specimens were recently found to perform similarly to clinician collected samples[**32**]. Moreover, there was no need for the clinical teams to use additional PPE as they were already wearing it to provide dental care; and the patients could attend for their treatment despite periods of lockdown restriction.

Although large numbers of dental patients were recruited, the number of participants with positive test outcomes was too small to test for associations between positivity and demographic characteristics. However, there were sufficient numbers to monitor the trends of SARS-CoV-2 in the asymptomatic population over time. This dental surveillance program is continuing through the winter and is now complementing the larger ONS survey which is currently being rolled out across Scotland.

## Conclusions

To our knowledge this is the first COVID-19 surveillance survey in dental settings, and we have demonstrated the feasibility of developing and implementing a surveillance testing protocol at a rapid pace in response to the pandemic. Participating patients were largely representative of the Scottish population. The positivity rate in this patient group reflects the underlying prevalence in the community at the time.

Our data have contributed to Public Health Scotland’s enhanced surveillance work, which is monitoring how COVID-19 is spreading through the population of Scotland via collation of a wide variety of data about COVID-19 from a range of sources. As the pandemic has evolved, this surveillance work has supported the Scottish Government with the national response to the pandemic.

These data are a salient reminder, particularly when community infection levels are rising, of the importance of appropriate ongoing Infection Prevention Control and PPE vigilance, which is relevant as healthcare team fatigue increases as the pandemic continues.

Dental settings are a valuable location for public health surveillance.

## Data Availability

Data are available on application to Electronic Data Research and Innovation Service (eDRIS) Public Health Scotland.

https://www.isdscotland.org/products-and-services/edris/

## Author Contributions

DC, SC, ME Contributed to conception, design, data acquisition and interpretation, drafted and critically revised the manuscript

CC, CW, CR Contributed to data acquisition and statistical analysis

RB, EO’K, NMcG, JB, SP Contributed to design, data acquisition and interpretation. HV, HJG, RG contributed to data acquisition and interpretation.

All authors gave their final approval and agree to be accountable for all aspects of the work.

## Acknowledgements

**\* Dental COVID-19 Surveillance Group**

We are grateful to the support of many people many dental and public health teams across Scotland for their contributions to the COVID-19 surveillance effort in Scotland (up to end November 2020). The people listed organised sites and logistics for swab collection and local data collection. We sincerely thank the dental teams who supported all the individuals listed below. **NHS Greater Glasgow & Clyde:** Lee Savarrio, Michael McGrady, Clare Brown, Julie Campbell, Lisa Harrigan, Linda McGrath, Karen Gallacher, Susan Frew, Shona Reid, Callum Wemyss, Simon Hobson, Jill Sweeney, Pei Rong Rong Chua, Siti Binti Mohd Khairi; **NHS Fife**, Dawn Adams, Debbie Slidders, Pamela Balfour, Catherine Balfour, Claire Connor; **NHS Dumfries & Galloway:** Alison Milne, Valerie White, Vicki Rice; **NHS Lothian:** Angus Walls, Christopher Carter, Lisa Anderson, Alex Keir, Christine Peaker, Cathie Gunn, Jane Locke, Dominika Janicka, Kenny Elliot; **NHS Tayside:** Morag Curnow, Susan Carson, Thomas Richmond, Hal Esler, Christopher Wright, Kathryn Walker, Heather Lackie, Fiona Fenton, Iain Hay, Claire Ryan; **NHS Borders:** Morag McQuade, Susan Carson, Sarah Brodie, Aileen Richardson, Pauline Hogg, Nicola Beattie, Louise Ponton; **NHS Highland:** John Lyon, Kirstin Edmiston, Nicola Anderson; **NHS Ayrshire & Arran:** Peter Ommer, June Johnston, Lisa Neill, Karen Parker, Tracy Pryce, Janet Campbell, Amanda Crombie, Diane Hunter, Morag Muir; **NHS Forth Valley:** Jennifer Rodgers, Susan Carson, Lesley Yeaman, Lynn Shearer, Gordon Morson, Joanne Conlin, Fiona MacPhail; **NHS Lanarkshire:** Anne Moore, Albert Yeung, Jean Kerr, Alison Kerr, Laura Milby, Shelley Percival; **NHS Western Isles:** Colin Robertson, Amanda MacKay, Eric MacDonald; **NHS Grampian:** Jonathan Iloya, Carol Craig, Elaine Dey, Laura Anderson, Mike Brown, Joanne Gallagher, Amy Ward; **NHS Shetland:** Antony Visocchi, Morag Mouat, Rachel Hunter, Gwen Farmer; **NHS Orkney:** Jay Wragg, Jennifer Watt. We extend our thanks to the Health Board Public Health / Health Protection and Test & Protect teams. And in the **UKGOV/DHSC Lighthouse Laboratory network:** Anna Dominiczak, Carol Clugston, Claire Churchill, Rianna Poulos, Sarah Hall, Lara Wigdor. In **Public Health Scotland** we acknowledge colleagues from across the organisation for their support including Megan Gorman, Danny Nicholson, Suzanne Redmond, Pam McVeigh, Debbie Sigerson, Julie Drysdale, Amanda Burridge, Josie Murray, David Goldberg. We also thank **The Scottish Government** for their support including Tom Ferris, Hugh McAloon, Mary Morgan, Janis Heaney.

Finally, we wish to thank all the dental patients for agreeing to participate.

---

**Appendix Table 1.** Positive test Cycle Threshold (CT) values and interpretation.

| Positive Test | CT Values and Interpretation |  |  |  |  |  |  |  |  |
| --- | --- | --- | --- | --- | --- | --- | --- | --- | --- |
|  | CH1-Target - ORF1ab |  | CH2-Target - N Gene |  | CH3-Target - S Gene |  | CH4-Target - MS2 |  | Interpretation |
|  | CH1-Result | CH1-Cq | CH2-Result | CH2-Cq | CH3-Result | CH3-Cq | CH4-Result | CH4-Cq |  |
| 1 | Positive | 19.47 | Positive | 21.54 | Positive | 21.76 | Positive | 22.11 | three gene positive |
| 2 | Positive | 21.64 | Positive | 22.91 | Positive | 22.1 | Positive | 24.15 | three gene positive |
| 3 | Positive | 29.61 | Positive | 30.45 | Positive | 30.17 | Positive | 22.19 | three gene positive weak/close to the limit of detection |
| 4 | Positive | 28.15 | Positive | 29.88 | Positive | 30.18 | Positive | 22.64 | three gene positive |
| 5 | Positive | 26.13 | Positive | 26.09 | Positive | 26.11 | Positive | 21.34 | three gene positive |
| 6 | Positive | 28.39 | Positive | 29.11 | Positive | 28.95 | Positive | 21.09 | three gene positive |
| 7 | Positive | 15.5 | Positive | 17.66 | Positive | 18.25 | Positive | 25.33 | three gene positive |
| 8 | Positive | 32.67 | Negative | ND | Negative | ND | Positive | 20.69 | single gene weak/close to the limit of detection |
| 9 | Positive | 22.58 | Positive | 24.72 | Positive | 23.27 | Positive | 22.93 | three gene positive |
| 10 | Positive | 20.98 | Positive | 21.87 | Positive | 21.71 | Positive | 22.31 | three gene positive |
| 11 | Positive | 35.83 | Negative | ND | Negative | ND | Positive | 22.98 | single gene weak/close to the limit of detection |
| 12 | Positive | 28.4 | Positive | 30.74 | Positive | 34.85 | Positive | 21.73 | three gene positive weak/close to the limit of detection |
| 13 | Positive | 30.7 | Positive | 30.73 | Positive | 31.71 | Positive | 20.83 | three gene positive weak/close to the limit of detection |
| 14 | Negative | ND | Positive | 34.47 | Negative | ND | Positive | 20.17 | single gene weak/close to the limit of detection |
| 15 | Positive | 31.28 | Positive | 32.32 | Negative | ND | Positive | 21.1 | two gene weak/close to the limit of detection |
| 16 | Negative | ND | Positive | 34.47 | Negative | ND | Positive | 20.11 | single gene weak/close to the limit of detection |
| 17 | Positive | 14.9 | Positive | 15.76 | Positive | 18.17 | Positive | 21.38 | three gene positive |
| 18 | Positive | 26.72 | Positive | 25.75 | Positive | 25.67 | Positive | 20.51 | three gene positive |
| 19 | Positive | 28.62 | Positive | 29.39 | Positive | 27.72 | Positive | 21.23 | three gene positive |
| 20 | Negative | ND | Positive | 31.7 | Positive | 33.12 | Positive | 21.05 | two gene weak/close to the limit of detection |
| 21 | Positive | 32.8 | Negative | ND | Negative | ND | Positive | 20.71 | single gene weak/close to the limit of detection |
| 22 | Negative | ND | Positive | 34.28 | Negative | ND | Positive | 21.89 | single gene weak/close to the limit of detection |

CT – Cycle Threshold; ND – Not Detected

